# The Genetic Risk of Gestational Diabetes in South Asian women

**DOI:** 10.1101/2022.07.13.22277599

**Authors:** Amel Lamri, Jayneel Limbachia, Karleen M Schulze, Dipika Desai, Brian Kelly, Russell J de Souza, Guillaume Paré, Deborah A Lawlor, John Wright, Sonia S Anand, the Born in Bradford and START investigators

**Author notes:** ***Corresponding Author:*** Dr. Sonia S Anand, McMaster University, MDCL 3202, 1280 Main St W, Hamilton, Ontario L8S 4K1, Canada. **e-mail:**.

## Abstract

South Asian women are at increased risk of developing gestational diabetes (GDM). Few studies have investigated the genetic contributions to GDM risk. We investigated the association of a type 2 diabetes (T2D) polygenic risk score (PRS), on its own, and with GDM risk factors, on GDM-related traits using data from two birth cohorts in which South Asian women were enrolled during pregnancy. 837 and 4,372 pregnant South Asian women from the SouTh Asian BiRth CohorT (START) and Born in Bradford (BiB) cohort studies underwent a 75-gram glucose tolerance test. PRSs were derived using GWAS results from an independent multi-ethnic study (∼18% South Asians). Associations with fasting plasma glucose (FPG); 2h post-load glucose (2hG); area under the curve glucose; and GDM were tested using linear and logistic regressions. The population attributable fraction (PAF) of the PRS was calculated. Every 1 SD increase in the PRS was associated with a 0.085 mmol/L increase in FPG ([95%CI=0.07-0.10], P=2.85 × 10^−20^); 0.21 mmol/L increase in 2hG ([95%CI=0.16-0.26], P=5.49 × 10^−16^); and a 45% increase in the risk of GDM ([95%CI=32-60%], P=2.27 × 10^−14^), independent of parental history of diabetes and other GDM risk factors. PRS tertile 3 accounted for 12.5% of the population’s GDM. No consistent interactions of the PRS with BMI, or diet quality were observed. A T2D PRS is strongly associated with multiple GDM-related traits in women of South Asian descent, and accounts for a substantial proportion of the PAF of GDM.

## INTRODUCTION

Gestational diabetes mellitus (GDM) is defined as hyperglycaemia first diagnosed during pregnancy. This abnormal increase in blood glucose levels is associated with an increased risk of adverse health outcomes for both mother and their fetus/child during pregnancy, and later in life.(Farrar et al., 2016) It is estimated that 1% to >30% of live births are affected by GDM worldwide. This prevalence has been shown to vary widely depending on the participants ethnicity, countries/regions, and on the diagnostic criteria used.(Archambault, Arel, & Filion, 2014; McIntyre et al., 2019) South Asian women (whose ancestry derives from the Indian subcontinent) have a 2-fold increased odds of developing GDM, compared to white European women.(Anand et al., 2016; Cosson et al., 2014; Farrar et al., 2015; McIntyre et al., 2019) The reasons for this disproportionate risk have not been fully characterized.

Gestational diabetes is a complex disorder influenced by multiple genetic and environmental factors such as maternal age, ethnicity, obesity, poor diet quality, and family history of diabetes.(Anand et al., 2017; Hedderson, Darbinian, Quesenberry, & Ferrara, 2011; Solomon et al., 1997) Most genetic and environmental GDM risk factors are shared with type 2 diabetes, (Sattar & Greer, 2002; Zhang & Ning, 2011) another condition which is thought to be very closely related to GDM. For example, women with GDM have a higher probability of having at least one parent with type 2 diabetes, compared to those with normal gestational glycemia.(Jang, Min, Lee, Cho, & Metzger, 1998) Furthermore, women with a GDM history have a 10-fold higher risk of subsequently being diagnosed with type 2 diabetes compared to those without a history of GDM.(Vounzoulaki et al., 2020) In terms of genetic architecture, both candidate gene and genome-wide association studies (GWASs) demonstrated a considerable overlap between GDM and type 2 diabetes.(Hayes et al., 2013; Kwak et al., 2012; Pervjakova et al., 2021) Finally, type 2 diabetes polygenic risk scores (PRSs) have also been associated with GDM risk.(Lamri et al., 2020; Pervjakova et al., 2021)

It has been demonstrated that environmental exposures such as diet and/or physical activity may modulate the effect of type 2 diabetes loci (such as *TCF7L2, PPARG* and *CDKAL1*) on the risk of type 2 diabetes.(Dietrich et al., 2019) Nevertheless, only a handful of studies have investigated genetic×environmental interactions on GDM,(Chen et al., 2019; Grotenfelt et al., 2016; Popova et al., 2017) and to date, no study has tested the interaction between a genome-wide PRS with other GDM risk factors, on the risk of GDM.

The aims of this investigation were to: *i*) test the association of a type 2 diabetes PRS, generated from an external multi-ethnic GWAS (∼18% South Asians), with GDM and related traits (fasting plasma glucose (FPG), 2h post-load glucose (2hG) and area under the curve glucose (AUCg) levels) in pregnant South Asian women from the SouTh Asian biRth cohorT (START) and the Born in Bradford (BiB) studies; *ii*) To estimate the population attributable fraction (PAF) of the PRS on GDM, and *iii*) To determine whether the effect of the PRS is modulated by other GDM risk factors including age, BMI, diet quality, birth country, education, and parity.

## METHODS

### Study design and participants

START is a prospective cohort study designed to evaluate the environmental and genetic determinants of cardio-metabolic traits among South Asian women and their offspring living in Canada. (Anand et al., 2013) In brief, 1,012 South Asian pregnant women, aged between 18 and 40 years old, were recruited during their second trimester of pregnancy from the Peel Region (Ontario, Canada) through physician referrals between 2011 and 2015. All START participants provided informed consent, and the study was approved by local ethics committees (Hamilton Integrated Research Ethics Board [ID:10-640], William Osler Health System [ID:11-0001], and Trillium Health Partners [RCC:11-018, ID:492]).

BiB is a prospective, longitudinal family cohort study designed to investigate the causes of illness, and develop interventions to improve health in a deprived multi-ethnic population in Bradford, England, UK.(Wright et al., 2013) Between 2007 and 2011, 12,453 women of various ethnic backgrounds (∼46% South Asian origin) were recruited between their 24^th^ and 28^th^ week of pregnancy. Detailed information on socio-economic characteristics, ethnicity, family history, environmental and physical risk factors has been collected.(Farrar et al., 2015; Wright et al., 2013) Ethical approval for all aspects of the research was granted by Bradford Research Ethics Committee [Ref 07/H1302/112].

#### *Measurements and* questionnaires

##### START

A detailed description of the maternal measurements has been published previously.(Anand et al., 2017) Briefly, weight and height were measured using standard procedures, and information about pre-pregnancy weight, family and personal medical history was collected using questionnaires. Parental history of diabetes was derived from baseline questionnaires and categorized as neither parents had a history of diabetes or either one or both parents did. Birth country, number of years spent in Canada, and education-related variables were self-reported. Participants’ highest level of education was coded as a five-category ordinal variable as: 1) less than high school; 2) high school completed; 3) Diploma or certificate from trade, technical or vocational school; 4) Bachelor’s or undergraduate degree, or teacher’s college; 5) Master’s, Doctorate or professional degree. A binary “born in South Asia” variable was categorized as participants born in South Asia (India, Pakistan, Sri Lanka or Bangladesh versus participants were born in any other country. A validated ethnic-specific food frequency questionnaire (FFQ) was used to collect dietary information.(Kelemen et al., 2003) To calculate diet quality, 1 point was given for consuming ≥ the study population median of: 1) green leafy vegetables, 2) raw vegetables, 3) other cooked vegetables, and 4) fruits, and less than the population median of 5) fried foods/fast food/ or snacks, and 6) meat/poultry. The continuous scores (ranging from 0 to 6 points) was categorized into a binary variable: low diet quality vs. medium or high quality] before analysis. (Anand et al., 2017)

#### BiB

Maternal height was measured during the recruitment visit (24-28^th^ weeks of pregnancy) using standard procedures. In the absence of pre-pregnancy weight data, weight from the first antenatal clinic visit (average 12 weeks of pregnancy) was used to calculate BMI. Ethnicity of participants and years spent in UK were self-reported at recruitment through an interview administered questionnaire; missing ethnicity data was backfilled from primary care data when available. The South Asian ethnicity of all participants included in this analysis was validated using genetic data. Parental history of diabetes and “born in South Asia” variables were derived from the baseline questionnaire data and coded as in START. Since only a very small proportion of BiB’s participants completed an FFQ that included information about fruits and vegetables intake, the diet quality score could not be derived in BiB. Data regarding the participant’s highest educational qualification was equalized (using UK standards) and recoded into the following categories: 1) less than 5 General Certificate of Secondary Education (GCSE) equivalent; 2) 5 GCSE equivalent; 3) A-level equivalent; 4) higher than A-level. Data for unclassifiable foreign degrees was considered as missing.

#### Outcomes

Study participants without prior type 2 diabetes were invited to undertake a 75-gram oral glucose tolerance test (OGTT) in both START and BiB, and FPG, and 2hG levels were measured (1h post-load glucose was measured in START only). AUCg was calculated using the FPG and 2hG glucose levels in BiB, and using the FPG, 1h post-load glucose, and 2hG levels in START.(Anand et al., 2017) Given the difference in the number of data points included in the calculation of AUC between the two studies and the skewness of the distributions, values were log-transformed, winsorized and standardized in each study before analysis. Gestational diabetes status of women without pre-existing type 2 diabetes was primarily defined based on OGTT results in both studies using the International Association of Diabetes and Pregnancy Study Group (IADPSG) GDM criteria (FPG ≥5.1 mmol/L or higher, or a 1hG ≥10.8 or a 2hG ≥ 8.5 mmol/L or higher)(International Association of et al., 2010). Our secondary outcome was GDM using BiB’s South Asian specific definition (FPG of 5.2 mmol/L or higher, or a 2hG of 7.2 mmol/L or higher) (Farrar et al., 2015), which will be referred to as the South Asian-specific definition hereafter. Self-reported GDM status or data from the birth chart was used to determine GDM’s status if OGTT measures were unavailable (N=65 and 31 in START and BiB respectively). Women with pre-existing diabetes at baseline were not included in this analysis. Pre-pregnancy diabetes status was determined using maternal self-reported data (about diabetes diagnosis, diabetes medication and/or insulin intake prior to pregnancy) in START. In BiB, information on pre-pregnancy diabetes was backfilled from electronic medical records.

In order to keep a single pregnancy (and a single GDM status) per mother in BiB, only pregnancies with no missing data for GDM were included. For mothers with available data at multiple pregnancies at this stage, pregnancies with no missing data across all covariates (age, BMI, family history, birth country, parity, education level) were prioritized. Next, only pregnancies with the least amount of missing data across all covariates were kept. The following two additional filtering approaches were then applied for mothers with multiple pregnancies remaining: i) if GDM was not diagnosed at any of the pregnancies, phenotype data at the latest available time point was kept (ie. keep older GDM controls). ii) if GDM was diagnosed during any of the pregnancies included in the study, the earliest time point where GDM was diagnosed was kept (ie. keep younger GDM cases).

### DNA extraction, Genotyping, Imputation and Filtering

#### START

DNA was extracted and genotyped for 867 mothers using the Illumina Human CoreExome-24 and Infinium CoreExome-24 arrays (Illumina, San-Diego, CA, USA). 837 samples passed standard quality control procedures.(Anderson et al., 2010) Genotypes were phased and imputed using SHAPEIT v2.12(Delaneau, Marchini, Genomes Project, & Genomes Project, 2014), and IMPUTE v2.3.2(Howie, Donnelly, & Marchini, 2009) respectively using the 1000 Genomes (phase 3) data as a reference panel.(Consortium et al., 2015) Variants with an info score <0.7 were removed from analysis. In total, 837 START participants with both genotypes and available GDM status, FPG, 1h- and/or 2hG levels were included in the analysis (Figure S1).

#### Born in Bradford

DNA was extracted and genotyped for 16,267 and 3,663 BiB participants using the Illumina HumanCoreExome (12v1.0, 12v1.1 or 24v1.0) and InfiniumGlobal Screening Array (24v2.0) arrays respectively (Illumina, San-Diego, CA, USA). 4,372 South Asian mothers passed genotyping quality controls, had GDM status, FPG, and/or 2hG levels available, and were included in our analysis (Figure S1).

#### Deriving the PRS

Given the absence of publicly available South Asian-specific T2D or GDM GWAS data at the time of the analysis, weights were derived from the DIAGRAM’s 2014 multi-ethnic T2D GWAS meta-analysis, which included over 18% of South Asians (∼63% European and 19% other ethnic backgrounds).(Mahajan et al., 2014) A grid search approach was used to identify the optimal parameters (17 P-values tested, ranging from 5×10^−8^ to 1 with 0.1 increase; 4 heritability values tested: 0.023; 0.06; 0.08; 0.12). START and BiB genotypes were pooled. 70% of the samples’ data were used for training and 30% for validation (random sampling stratified by study) in order to minimize the impact of population stratification. The PRS was derived using LDpred2.(Prive, Arbel, & Vilhjalmsson, 2020) The best PRS (i.e. that maximized the AUC) was characterized by a P value ≤ 0.0014 and an h^2^=0.08 (N_SNVs_=6,492). The PRS was standardized (mean=0, standard deviation=1) in both studies before analysis.

#### Principal component analysis of genetic data

A principal component analysis (PCA) was performed using the PC-Air function from the GENESIS R package (v2.20.0).(Conomos, Miller, & Thornton, 2015) Kinship matrices (required to derive PCs with PC-Air) were derived using KING (v2.2.5).(Manichaikul et al., 2010)

### Statistical Analysis

The statistical analysis was conducted using R (v3.6.3).(R core Team, 2016) Linear regression models were used to test the association between the PRS and FPG, 2hG and AUCg. PRS and GDM associations were tested using logistic regression. Both univariate and multivariate models were constructed with adjustment for GDM risk factors (age, BMI, parity, birth in South Asia (yes *vs*. no), education level, and diet quality (in START only) and the first 5 PCs (in order to minimize the effect of population stratification). Interactions between the PRS and each risk factors was also tested. The estimated population attributable fractions (PAFs) and their corresponding standard errors were calculated using the AF R package (v.0.1.5). To this end, continuous variables were recoded into categorical variables: age was divided in two categories [(29-31, 32-43) vs. 19-28]; BMI was stratified into a two categories variable using South Asian obesity cutoff points suggested by Gray et al. (Gray et al., 2011) (<23 vs. ≥23); The PRS was divided into two categories (tertiles 1+2 vs tertile 3); Parity was divided into two categories (primiparity vs. 1 pregnancy or more); Education level variables were divided in two categories (completed high school or lower vs. higher degree, diploma or certificate in START; and A-level equivalent or lower vs. higher than A-level in BiB).

## RESULTS

The proportion of women classified with GDM using the IADPSG criteria was 25% and 11.2% in START and BiB respectively, which was lower than the proportion using the South Asian-specific definition of 36.2% and 22.9% respectively. Notably the proportion of women with GDM was higher in START compared to BiB irrespective of the classification method used.

The proportion of women of Indian origin in START and BiB was 71.8% and 5.1%, while the proportion of Pakistani women was 23.4% and 94.3% respectively. The proportion of participants born in the Indian sub-continent was higher in START (88.6%) than in BiB (55.6%), and the average number of years spent in Canada or the UK among these participants was lower in START compared to BiB (6.6 vs. 9.7 years respectively). The proportions of primiparous women (40.9% vs 31.7%) and women with 1 prior pregnancy (42.4% vs 26.9%) were higher in START than in BiB. Conversely, participants with 2 or more prior pregnancies were more frequent in BiB than START (41.4% vs. 16.6% respectively). The proportion of vegetarian participants was higher in START than in BiB (36.4% vs 1.3%). Finally, the proportion of participants with a post-secondary degree/diploma or higher was greater in START than BiB (84.0% vs 29.0%).

Table 1 shows the baseline characteristics of the South Asian women from the START and BiB stratified by GDM case vs non GDM (IADPSG criteria). As expected, women with GDM had a higher mean fasting, 2hG and AUCg levels than non-GDM participants. Participants with GDM were older, had a higher BMI, and were more likely to report a family history of diabetes compared to women without GDM, in both studies. The overall diet quality was lower in participants with GDM compared to non-GDM participants in START (data not available in BiB). Of note, the average difference in BMI between GDM cases and controls was higher in BiB than in START (2.8 and 1.8 respectively) (Table 1).

**Table 1:**
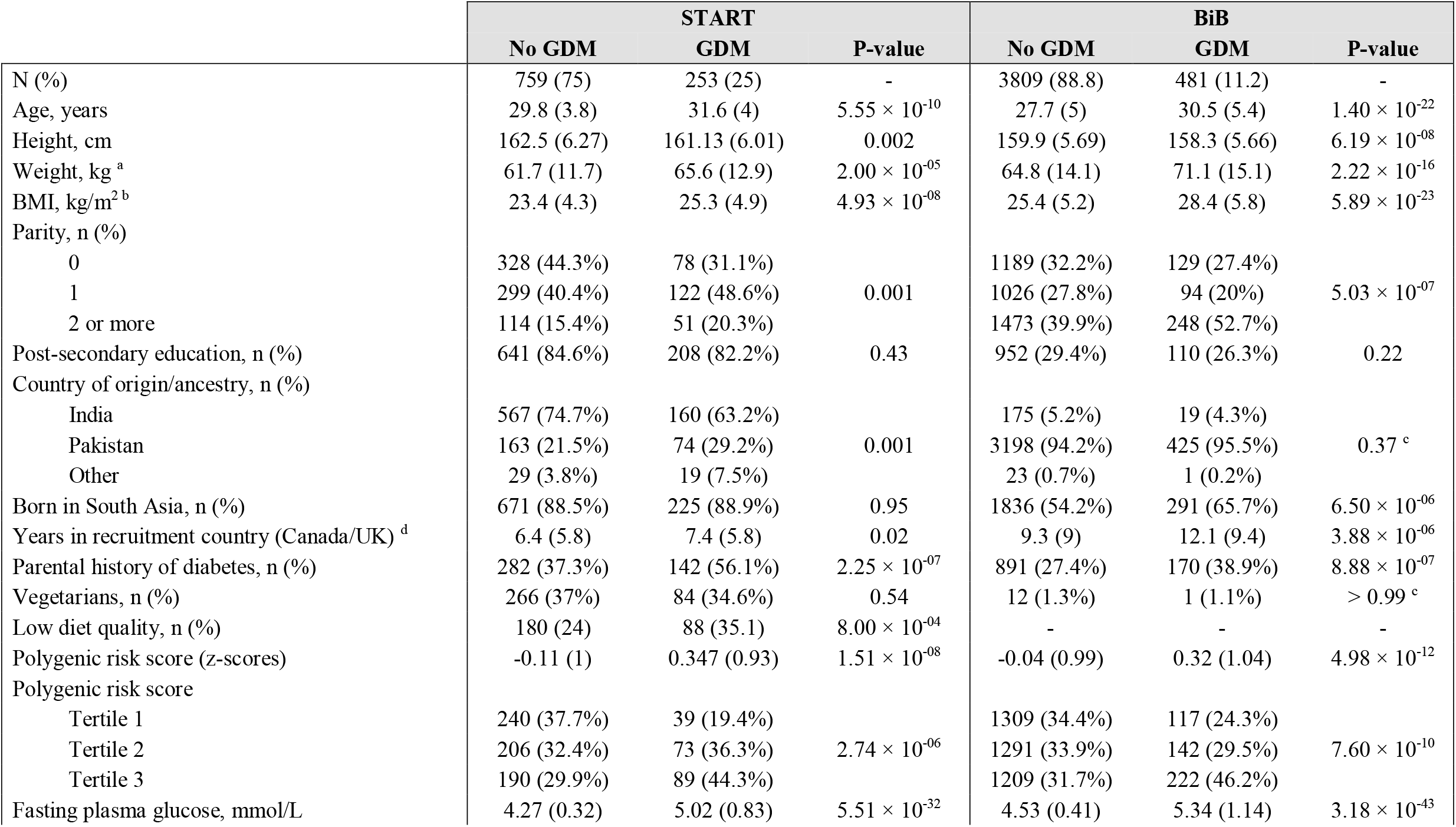
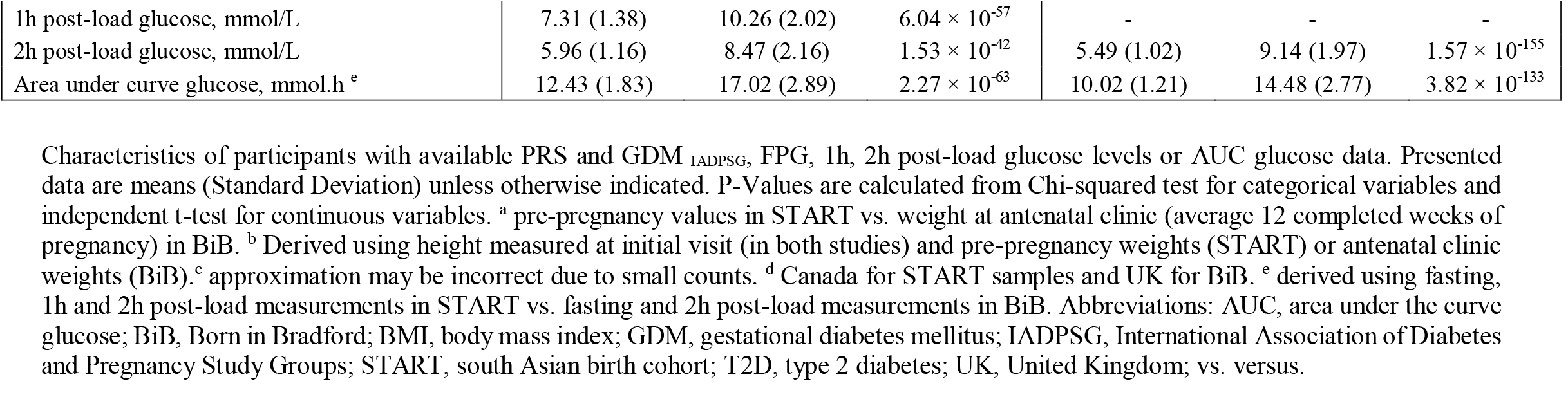
Characteristics of START and BiB study participants included in the analysis.

|  | START |  |  | BiB |  |  |
| --- | --- | --- | --- | --- | --- | --- |
|  | No GDM | GDM | P-value | No GDM | GDM | P-value |
| N (%) | 759 (75) | 253 (25) | - | 3809 (88.8) | 481 (11.2) | - |
| Age, years | 29.8 (3.8) | 31.6 (4) | $5.55 \times 10^{-10}$ | 27.7 (5) | 30.5 (5.4) | $1.40 \times 10^{-22}$ |
| Height, cm | 162.5 (6.27) | 161.13 (6.01) | 0.002 | 159.9 (5.69) | 158.3 (5.66) | $6.19 \times 10^{-08}$ |
| Weight, kg <sup>a</sup> | 61.7 (11.7) | 65.6 (12.9) | $2.00 \times 10^{-05}$ | 64.8 (14.1) | 71.1 (15.1) | $2.22 \times 10^{-16}$ |
| BMI, kg/m<sup>2</sup> <sup>b</sup> | 23.4 (4.3) | 25.3 (4.9) | $4.93 \times 10^{-08}$ | 25.4 (5.2) | 28.4 (5.8) | $5.89 \times 10^{-23}$ |
| Parity, n (%) |  |  |  |  |  |  |
| 0 | 328 (44.3%) | 78 (31.1%) |  | 1189 (32.2%) | 129 (27.4%) |  |
| 1 | 299 (40.4%) | 122 (48.6%) | 0.001 | 1026 (27.8%) | 94 (20%) | $5.03 \times 10^{-07}$ |
| 2 or more | 114 (15.4%) | 51 (20.3%) |  | 1473 (39.9%) | 248 (52.7%) |  |
| Post-secondary education, n (%) | 641 (84.6%) | 208 (82.2%) | 0.43 | 952 (29.4%) | 110 (26.3%) | 0.22 |
| Country of origin/ancestry, n (%) |  |  |  |  |  |  |
| India | 567 (74.7%) | 160 (63.2%) |  | 175 (5.2%) | 19 (4.3%) |  |
| Pakistan | 163 (21.5%) | 74 (29.2%) | 0.001 | 3198 (94.2%) | 425 (95.5%) | 0.37 <sup>c</sup> |
| Other | 29 (3.8%) | 19 (7.5%) |  | 23 (0.7%) | 1 (0.2%) |  |
| Born in South Asia, n (%) | 671 (88.5%) | 225 (88.9%) | 0.95 | 1836 (54.2%) | 291 (65.7%) | $6.50 \times 10^{-06}$ |
| Years in recruitment country (Canada/UK) <sup>d</sup> | 6.4 (5.8) | 7.4 (5.8) | 0.02 | 9.3 (9) | 12.1 (9.4) | $3.88 \times 10^{-06}$ |
| Parental history of diabetes, n (%) | 282 (37.3%) | 142 (56.1%) | $2.25 \times 10^{-07}$ | 891 (27.4%) | 170 (38.9%) | $8.88 \times 10^{-07}$ |
| Vegetarians, n (%) | 266 (37%) | 84 (34.6%) | 0.54 | 12 (1.3%) | 1 (1.1%) | $> 0.99$ <sup>c</sup> |
| Low diet quality, n (%) | 180 (24) | 88 (35.1) | $8.00 \times 10^{-04}$ | - | - | - |
| Polygenic risk score (z-scores) | -0.11 (1) | 0.347 (0.93) | $1.51 \times 10^{-08}$ | -0.04 (0.99) | 0.32 (1.04) | $4.98 \times 10^{-12}$ |
| Polygenic risk score |  |  |  |  |  |  |
| Tertile 1 | 240 (37.7%) | 39 (19.4%) |  | 1309 (34.4%) | 117 (24.3%) |  |
| Tertile 2 | 206 (32.4%) | 73 (36.3%) | $2.74 \times 10^{-06}$ | 1291 (33.9%) | 142 (29.5%) | $7.60 \times 10^{-10}$ |
| Tertile 3 | 190 (29.9%) | 89 (44.3%) |  | 1209 (31.7%) | 222 (46.2%) |  |
| Fasting plasma glucose, mmol/L | 4.27 (0.32) | 5.02 (0.83) | $5.51 \times 10^{-32}$ | 4.53 (0.41) | 5.34 (1.14) | $3.18 \times 10^{-43}$ |
| 1h post-load glucose, mmol/L | 7.31 (1.38) | 10.26 (2.02) | $6.04 \times 10^{-57}$ | - | - | - |
| 2h post-load glucose, mmol/L | 5.96 (1.16) | 8.47 (2.16) | $1.53 \times 10^{-42}$ | 5.49 (1.02) | 9.14 (1.97) | $1.57 \times 10^{-155}$ |
| Area under curve glucose, mmol.h <sup>e</sup> | 12.43 (1.83) | 17.02 (2.89) | $2.27 \times 10^{-63}$ | 10.02 (1.21) | 14.48 (2.77) | $3.82 \times 10^{-133}$ |

Characteristics of participants with available PRS and GDM<sub>IADPSG</sub>, FPG, 1h, 2h post-load glucose levels or AUC glucose data. Presented data are means (Standard Deviation) unless otherwise indicated. P-Values are calculated from Chi-squared test for categorical variables and independent t-test for continuous variables. <sup>a</sup> pre-pregnancy values in START vs. weight at antenatal clinic (average 12 completed weeks of pregnancy) in BiB. <sup>b</sup> Derived using height measured at initial visit (in both studies) and pre-pregnancy weights (START) or antenatal clinic weights (BiB). <sup>c</sup> approximation may be incorrect due to small counts. <sup>d</sup> Canada for START samples and UK for BiB. <sup>e</sup> derived using fasting, 1h and 2h post-load measurements in START vs. fasting and 2h post-load measurements in BiB. Abbreviations: AUC, area under the curve glucose; BiB, Born in Bradford; BMI, body mass index; GDM, gestational diabetes mellitus; IADPSG, International Association of Diabetes and Pregnancy Study Groups; START, south Asian birth cohort; T2D, type 2 diabetes; UK, United Kingdom; vs. versus.

The standardized PRS ranged between −3.23 and 3.12 in START as compared to −3.51 and 4.16 in BiB. The full list of genetic variants included in the PRS as well as their characteristics are shown in Table S1. Women with GDM had a higher mean PRS compared to women without GDM. Similarly, women with GDM were more likely to have PRS categorized in tertile 2 or 3, compared to tertile 1 (Table 1).

### Genetic risk and GDM related traits in univariate models

The continuous PRS was associated with FPG, 2hG, and AUCg in START and BiB in univariate models. Every 1 SD increase in the PRS was associated with a 0.09 mmol/L increase in FPG [95%CI=0.07-0.10], 0.23 mmol/L increase in 2hG [95%CI=0.18-0.28], and a 0.17 unit increase in AUCg z-scores [0.14 - 0.20] in the meta-analysed results (Table S2).

The PRS was also associated with the risk of GDM _IADPSG_ in univariate models whereby a 1 SD increase in PRS was associated with a 47% increase in risk of GDM after meta-analysis [95%CI=35-60%]. A similar association is observed using the South Asian-specific definition of GDM, with moderate between-study heterogeneity observed (Table S2).

Overall, the risk of GDM _IADPSG_ increased progressively comparing tertile 2 of the PRS to tertile 1, and tertile 3 to tertile 1 (43% and 230% respectively, Table S2). Higher PRS categories were also associated with higher FPG, 2hG and AUCg levels (Table S2).

### Multivariable models of GDM risk factors and GDM related traits

The continuous PRS was strongly and independently associated with FPG, 2hG and AUCg levels in a multivariable model adjusted for age, BMI, parity, parental history of diabetes, region of birth (South Asia vs. other), education level, and diet quality (available in START only), and the first 5 PCs (Table 2). For example, every 1 SD increase in the PRS was associated with a 0.08 mmol/L increase in FPG, and 0.21 mmol/L increase in 2hG levels (Table 2). The continuous PRS was also associated with a higher risk of GDM in a model with similar adjustments whereby every 1 SD increase in the PRS was associated with a 45% increase in the risk of GDM _IADPSG_ (Table 2). Association results for GDM using the South Asian-specific criteria are shown in Table S3 When testing tertiles of PRS with similar covariates, our results show that participants in the 2^nd^ and 3^rd^ PRS tertiles have a 37% and 19% increase in the risk of GDM _IADPSG_ compared to participants in tertile 1 respectively (Table S4). Higher PRS tertiles were also associated with higher FPG, 2hG and AUCg levels (Table S4). The effect sizes associated with tertiles 2 were higher in START than BiB across multiple GDM related traits (2hG, AUCg and GDM, Table S4).

**Table 2:**
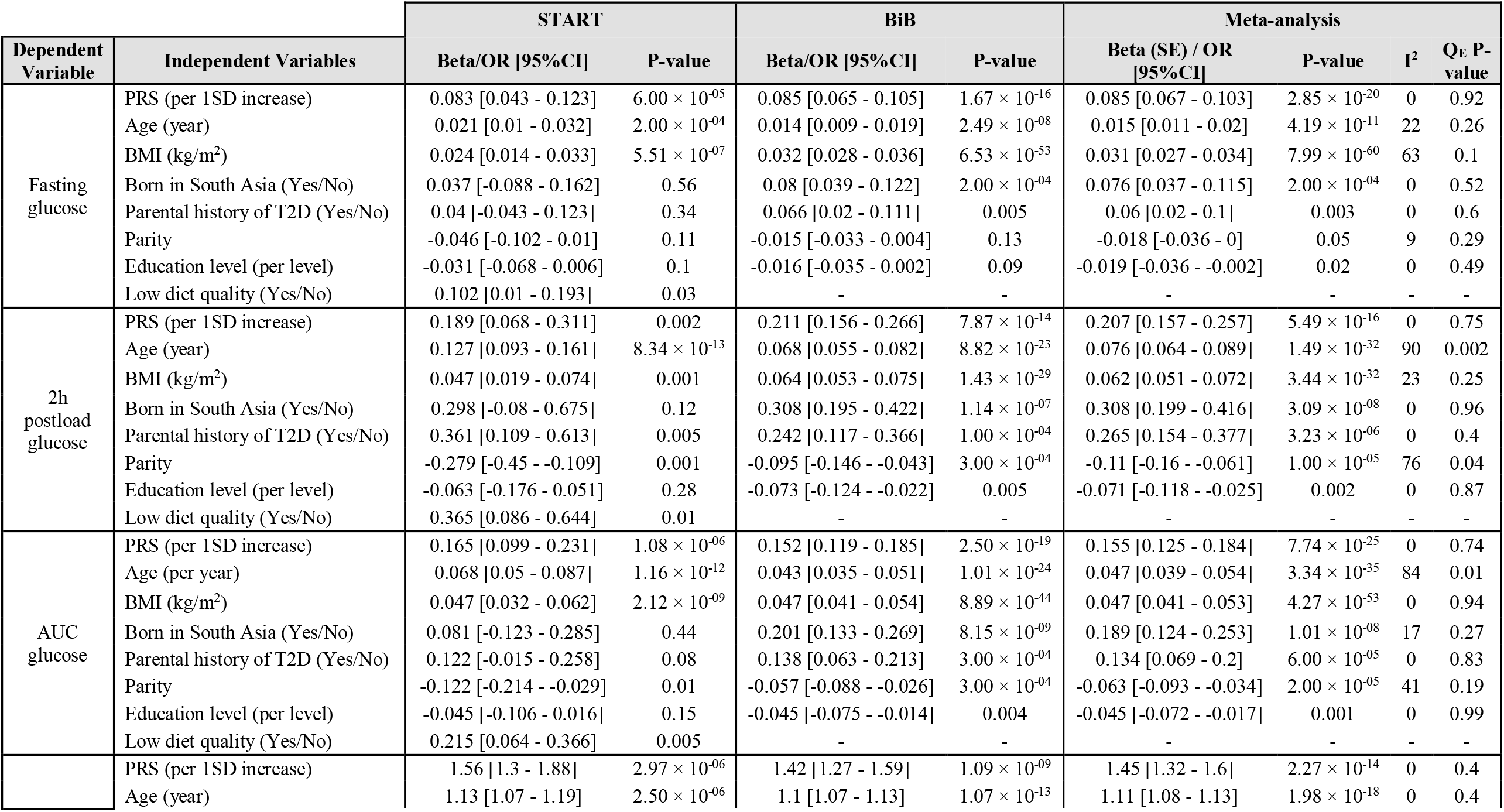
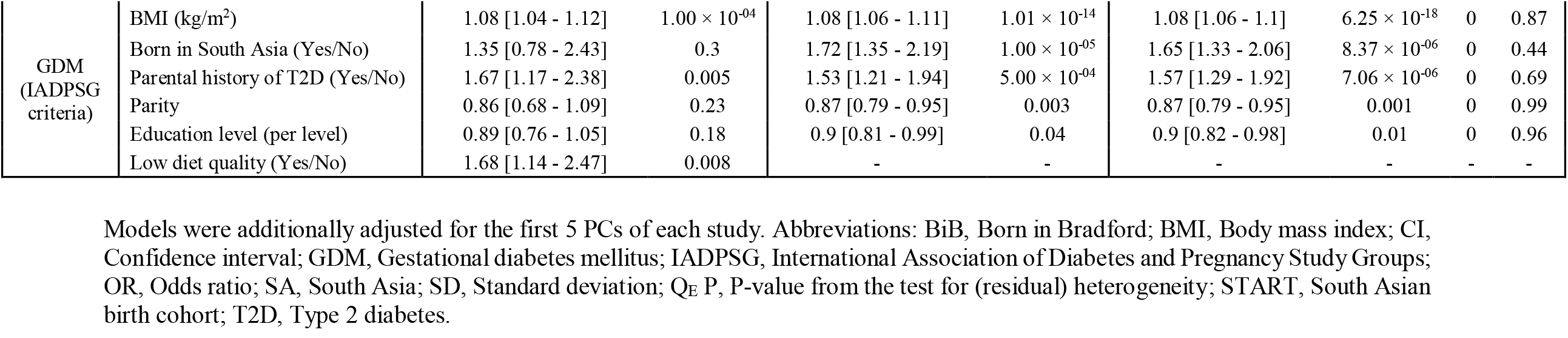
Association between GDM risk factors and GDM related traits: results from multivariate models in START and BiB cohorts.

### Population attributable fraction

In a model adjusted for maternal age, BMI, education, birth in South Asia (yes/no), parental history of diabetes, and diet quality (in START only), the PRS tertile 3 accounted for 12.5% of the population’s total GDM _IADPSG_ cases overall, and was higher in START than in BiB (Table 3). The combined effect of PRS and parental history of diabetes on GDM accounted for ∼21.7% of the population’s GDM cases in the two studies combined (Table 3).

**Table 3:** Population attributable fractions of GDM risk factors in mothers from the START and Born in Bradford studies (multivariable models).

| Independent Variable | START |  | BiB |  | Meta-analysis |  |  |  |
| --- | --- | --- | --- | --- | --- | --- | --- | --- |
|  | AF [95%CI] | P-value | AF [95%CI] | P-value | AF [95%CI] | P-value | I<sup>2</sup> | Q<sub>E</sub> P-value |
| Age (29-31 vs. <29 years) | 5.6 [-9.1 - 20.2] | 0.46 | 8.3 [3.5 - 13] | $6.00 \times 10^{-04}$ | 8 [3.5 - 12.5] | $5.00 \times 10^{-04}$ | 0 | 0.73 |
| Age (>32yr vs. <29 years) | 31.2 [17.1 - 45.3] | $1.00 \times 10^{-05}$ | 20.2 [14.8 - 25.7] | $4.72 \times 10^{-13}$ | 21.7 [16.6 - 26.8] | $9.19 \times 10^{-17}$ | 50 | 0.16 |
| Body mass index ($\geq 23$ vs. < 23) | 21.8 [8.7 - 34.9] | 0.001 | 33.8 [25.4 - 42.2] | $2.47 \times 10^{-15}$ | 30.3 [23.3 - 37.4] | $3.59 \times 10^{-17}$ | 56 | 0.13 |
| Born in SA (Yes vs. No) | 13.5 [-17.2 - 44.3] | 0.39 | 19.3 [12.6 - 26] | $1.47 \times 10^{-08}$ | 19 [12.5 - 25.6] | $1.07 \times 10^{-08}$ | 0 | 0.72 |
| Education (Post-secondary vs. less) | -18.2 [-46.8 - 10.5] | 0.21 | -0.8 [-4.6 - 3.1] | 0.7 | -1.1 [-4.9 - 2.7] | 0.58 | 28 | 0.24 |
| Parental history of T2D (Yes vs. No) | 15.1 [4.4 - 25.7] | 0.005 | 8.3 [4.1 - 12.5] | $1.00 \times 10^{-04}$ | 9.2 [5.3 - 13.1] | $3.54 \times 10^{-06}$ | 26 | 0.24 |
| PRS (Tertile 3 vs. 1+2) | 13.8 [4.9 - 22.6] | 0.002 | 12.2 [7.8 - 16.6] | $5.14 \times 10^{-08}$ | 12.5 [8.6 - 16.5] | $4.47 \times 10^{-10}$ | 0 | 0.76 |
| Low Diet Quality (Yes vs. No) | 8.9 [1.5 - 16.4] | 0.02 | - | - | - | - | - | - |
| Sum PAF of PRS (T3) and parental history of diabetes | 28.9 |  | 20.5 |  | 21.7 |  |  |  |

GDM status derived using IADPSG criteria. Multivariate models included age, BMI, region of birth (South Asia vs other), education, parental history of diabetes, parity, principal components 1 to 5, and diet quality (START only) when applicable. Abbreviations: BiB, Born in Bradford; BMI, Body mass index, CI, Confidence interval; GDM, Gestational diabetes mellitus; IADPSG, International Association of Diabetes and Pregnancy Study Groups; PAF, Population attributable fraction; PRS, Polygenic risk score; Q<sub>E</sub> P, P-value from the test for (residual) heterogeneity; START, South Asian birth cohort.

### Interactions between the PRS and GDM risk factors on GDM

No consistent interactions were observed between the PRS and maternal age; parity; or education level modulating FPG, 2hG, AUCg, or GDM _IADPSG_ in START or BiB (Table 4 and Table S5).

**Table 4:**
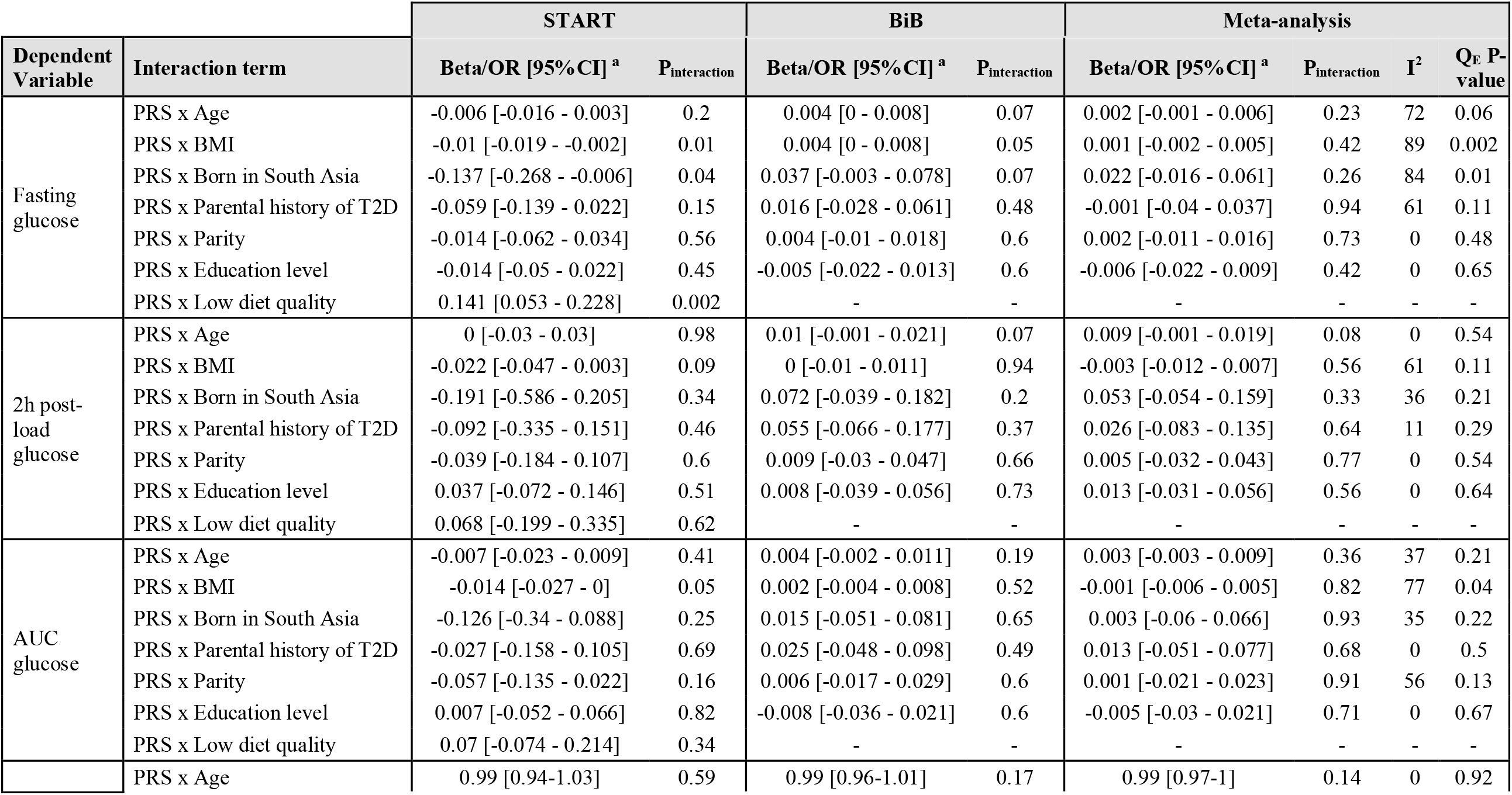
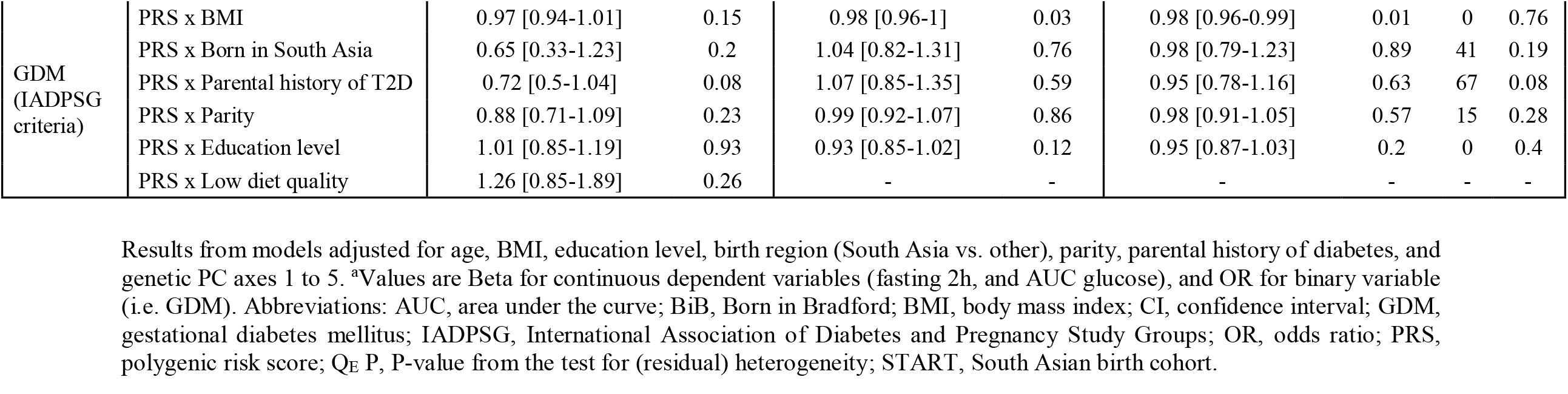
Interaction effects between GDM risk factors and T2D PRS in START and BiB.

Some nominally significant interactions modulating the continuous trait of FPG were observed in START which were not confirmed in BiB. These included the PRS with BMI and the PRS×birth in South Asia (yes/no) interactions (P_interaction_=0.01 and 0.04 respectively), yet non-significant in BiB (P_interaction_=0.05 and 0.07 respectively), with different effect sizes between the two studies (Table S6), resulting in non-significant meta-analysis of these effects (P_interaction_=0.45 and 0.26 respectively). Lastly, a PRS×diet quality interaction on FPG was detected in START (P_interaction_=0.002, Table 4) whereby the effect of the PRS appeared to be stronger in participants with a low diet quality (Beta=0.17 [95%CI=0.10-0.24]) than in participants with a medium or high diet quality (Beta=0.05 [95%CI=0.00-0.09]) (Table S6 and Figure S2). The overall diet quality score was not available BiB, hence this interaction could not be tested for replication.

## DISCUSSION

We demonstrate that a type 2 diabetes polygenic risk score, based on an independent and multi-ethnic GWAS meta-analysis (with 18% South Asian participants), is strongly associated with GDM and related glucose traits among South Asian pregnant women settled in Canada or the UK. This association is independent of other known GDM risk factors, including maternal age, BMI, parental history of diabetes, and maternal birth country. The PRS highest tertile accounted for 13.8 % of the PAF of GDM. Consistent with a recent trans-ethnicity GWAS of GDM, and these results support the hypothesis that GDM and type 2 diabetes are part of the same underlying pathology.(Pervjakova et al., 2021)

Family history of type 2 diabetes is often used as a surrogate marker of the genetic risk of type 2 diabetes. Our results show that the addition of the PRS to the multivariate models does not nullify the impact of parental history on GDM and *vice versa*. This suggests that the PRS and family history of diabetes both partially convey independent information. This partial independence could be explained by the fact that the PRS does not entirely capture the genetic association signals with GDM. On the other hand, family history reflects not only genetic similarity, but also shared non-genetic lifestyle factors.

Overall, we found no robust evidence for modulation of the PRS’s effect on GDM-related traits by other GDM risk factors. While marginal PRS×BMI and PRS×South Asia born interactions on FPG were observed, these did not replicate, both in terms of statistical significance and effect sizes, suggesting that these environmental and genetic factors may act independently. Furthermore, these interactions would not pass multiple testing corrections if applied. A potentially stronger PRS×diet quality interaction modulating FPG was observed in START. However, since it was not possible to replicate this interaction in BiB, future investigations are required in order to validate this observation. If confirmed, this interaction may help identify a subpopulation who will benefit the most from a targeted diet intervention for the prevention of GDM.

The overall clinical implications of our findings should be carefully considered. At present, the use of laboratory-derived genetic information in the clinical setting remains expensive and is not implemented for complex diseases like GDM. Given the enhanced predictive value of our genome-wide PRS, future evaluations of whether the knowledge of one’s genetic risk improves adherence to lifestyle recommendations and contributes to reducing gestational hyperglycemia would be of great interest.

Our study has been considerably strengthened by the use of a PRS optimized for a large population of South Asians from two independent cohorts, as well as by the fact that GDM status was determined using objective OGTT measures. Nevertheless, there are some limitations to our analysis that should be considered: *i)* The weights attributed to the genetic variants included in the PRS are derived from a type 2 diabetes study. Overall, evidence points to a strong correlation between top variants from type 2 diabetes and GDM GWASs. However, variants at some common loci (eg. *MTNR1B)* might have significantly different effect size depending on the phenotype studied.(Pervjakova et al., 2021) In addition, variants in at least one locus (*HKDC1)* have been strongly associated to GDM but not type 2 diabetes.(Pervjakova et al., 2021) More GDM-specific loci, or loci with a different magnitude of effect between GDM and type 2 diabetes might be identified from future, larger studies. These observations suggest that future PRSs based on a GDM GWAS may have a higher predictive value and more power to detect gene×environment interactions. *ii)* Second, some differences in measurements exist between START and BiB studies, including the timing of weight measurements, and the number of data points included in the calculation of AUCg. However, since data was standardized in both studies, we do not expect that AUCg measurements differences had a major impact on the results. *iii)* Finally, the comparison of genetic data between START and BiB revealed the existence of genetic heterogeneity, both between and within the samples of these two cohorts (Figure S3). It is our assumption that these differences can be explained by the difference of sample size (START being smaller than BiB), as well as by historical differences in migration patterns from South Asia to Canada and the UK. For example, most START participants were first generation migrants from India, whereas the majority of South Asians in BiB are descendants of Pakistani migrants who settled in the UK for several generations. In order to account for this genetic heterogeneity, we derived a new type 2 diabetes PRS that combined samples from the two studies. This PRS should be more generalizable to other South Asian studies. Another measure implemented to reduce the effect of population stratification was the adjustment for the PC axes in our analysis. Given the absence of heterogeneity in our FPG, 2hG, or GDM_IADPSG_ PC adjusted models, we consider that population stratification effects have been accounted for.

### Conclusion

A type 2 diabetes derived PRS is strongly associated with the risk of GDM in pregnant women of South Asian descent, independent of parental history of diabetes, and other GDM risk factors.

## Data Availability

: SSA, and JW hold the generated/analyzed START and BiB data used in this study respectively. The Data from START contains identifiable information that cannot be given to an outside group and is not publicly available as per the study's ethics guidelines. Nevertheless, requests for collaboration will be considered. Access to de-identified BiB data can be requested by following the instructions detailed here: https://borninbradford.nhs.uk/research/how-to-access-data/.

## Acknowledgements

Research projects in START and Born in Bradford are only possible because of the enthusiasm and commitment of the parents and children involved in these two studies. We are grateful to all the participants, teachers, school staff, health professionals and researchers and other contributors who have made these studies happen.

## Author contributions

**A.L**. derived the PRS, performed the association and interaction analysis, drafted and revised the manuscript. **J.L**. performed the association tests and PAR analysis in START, drafted and revised the manuscript. **K.S**. reviewed and supervised the statistical analysis and commented on the paper. **D.D**. is the study coordinator for the START birth cohort and provided comments on the manuscript. **B.K**. directed BiB data acquisition, provided comments on statistical analysis and reviewed the manuscript, **R.J.D**. provided comments on statistical analysis and reviewed manuscript. **G.P**. performed the genotyping laboratory analysis and provided comments on the manuscript. **D.A.L**. provided comments on the analysis and reviewed the manuscript. J.R. the BiB study’s principal investigator, he provided comments on statistical analysis and reviewed the manuscript. **S.S.A**. (guarantor) oversaw the development, analysis, and writing of this manuscript.

## Data and code availability

Data from START is not publicly available, since the study is bound by consent which indicates the data will not be used by an outside group. Requests for collaboration or replication will be considered for research purposes only (no commercial use allowed, as per the study’s informed consent). Requests should be addressed to the study’s principal investigator (Sonia Anand) via a form which will be provided upon request by emailing. The request will be evaluated by PIs and co-investigators, and projects deemed of scientific interest will be further evaluated/validated by local REB chair. Born in Bradford data are available for research purposes only by sending an expression of interest form downloadable from https://borninbradford.nhs.uk/wp-content/uploads/BiB_EoI_v3.1_10.05.21.doct to. The proposal will be reviewed by BiB’s executive team. If the request is approved, the requester will be asked to sign a <u>Data Sharing Contract</u> and a <u>Data Sharing Agreement</u>. Full details on how to access data and forms can be found here https://borninbradford.nhs.uk/research/how-to-access-data/. The code used to analyze the data is available at https://github.com/AmelLamri/Paper_T2dPrsGdm_StartBiB. All Sharable processed versions of the datasets used in the manuscript are made available as supplementary material or at https://github.com/AmelLamri/Paper_T2dPrsGdm_StartBiB.

## ABBREVIATIONS

1KG: 1000 Genomes
2hG: 2h post-load Glucose
AF: Attributable Fraction
AUC: Area Under the Curve
AUCg: AUC glucose
BiB: Born in Bradford
CI: Confidence Interval
DIAGRAM: DIAbetes Genetics Replication and Meta-analysis
DNA: DeoxyriboNucleic Acid
FFQ: Food Frequency Questionnaire
FPG: Fasting Plasma Glucose
GCSE: General Certificate of Secondary Education
GDM: Gestational Diabetes Mellitus
GWAS: Genome-Wide Association Study
IADPSG: International Association of Diabetes and Pregnancy Study Groups
OR: Odds Ratio
PAF: Population Attributable Fraction
PRS: Polygenic Risk Score
SE: Standard Error
START: SouTh Asian biRth cohort
T2D: Type 2 Diabetes
UK: United Kingdom

## URLs

1000 Genomes study

https://www.internationalgenome.org/

Born in Bradford Study

https://borninbradford.nhs.uk/

DIAGRAM consortium

https://www.diagram-consortium.org/

GCTA: a tool for Genome-wide Complex Trait Analysis

https://cnsgenomics.com/software/gcta

KING: Relationship Inference

https://people.virginia.edu/~wc9c/KING/manual.html

PC-AiR: Principal Components Analysis in Related Samples

https://rdrr.io/bioc/GENESIS/man/pcair.html

PLINK

https://www.cog-genomics.org/plink/

R Project

https://cran.r-project.org/

